# Modelling the interplay of SARS-CoV-2 variants in the United Kingdom

**DOI:** 10.1101/2021.11.26.21266485

**Authors:** N. L. Barreiro, T. Govezensky, C. I. Ventura, M. Núñez, P. G. Bolcatto, R. A. Barrio

## Abstract

Most COVID-19 vaccines have proved to be effective to combat the pandemic and to prevent severe disease but their distribution proceeds in a context of global vaccine shortage Their uneven distribution favors the appearance of new variants of concern, as the highly transmissible Delta variant, affecting especially non-vaccinated people. We consider that devising reliable models to analyse the spread of the different variants is crucial. These models should include the effects of vaccination as well as non-pharmaceutical measures used to contain the pandemic by modifying social behaviour. In this work, we present a stochastic geographical model that fulfills these requirements. It consists of an extended compartmental model that includes various strains and vaccination strategies, allowing to study the emergence and dynamics of the new COVID-19 variants. The models conveniently separates the parameters related to the disease from the ones related to social behavior and mobility restrictions. The geographical spread of the virus is modeled taking into account the actual population distribution in any given country of interest. Here we choose the UK as model system, taking advantage of the reliable available data, in order to fit the recurrence of the currently prevalent variants. Our computer simulations allow to describe some global features observed in the daily number of cases, as the appearance of periodic waves and the features that determine the prevalence of certain variants. They also provide useful predictions aiming to help planning future vaccination boosters. We stress that the model could be applied to any other country of interest.

## Introduction

The SARS-CoV-2 virus pandemic has infected more than 242 million people worldwide, with more than 4.9 million global deaths, by 22 Oct. 2021^1^. Global vaccination progressed this year, though at very different rates throughout the world and in a context of vaccine shortage. 48.3% of the world population received at least a first COVID-19 vaccine dose (in low-income countries this number drops to 3%)^2^. As the virus spreads, its genome replicates originating mutations that lead to virus variants.^3^

Several hundred thousand new variants have been identified in the SARS-CoV-2 genome^4^. Most of them have no effect on the transmissibility of the virus because they do not alter the “spike” protein shape^5^. Despite of this fact, the large amount of infections gave rise to some clinically relevant variants (variants of concern), that are transmitted more easily and rapidly than the original ones^6,7^ and that can even be more lethal^8^. The most widely spread strains include the ones first detected in: the United Kingdom (Alpha or B.1.1.7), South Africa (Beta or B.1.351), Brazil (Gamma or P.1), and India (Delta or B.1.617.2)^9^. The emergence of these new COVID-19 variants raises doubts about the effectiveness of vaccination campaigns^10^. Furthermore, these strains allowed the appearance of fast reinfections^11^ and co-infections^12^. The available information suggests that these new variants could draw out the pandemic if measures are not taken^13^. Further research is needed to understand the impact of their spread. Accordingly, we propose the development of a stochastic geographic expansion model, that includes several different strains, to study the emergence and dynamics of these new variants. The model is based on a previously used discrete mathematical map that starts from an extended SEIRS model^14–17^. The model is applied to the case of the UK by fitting the actual properties of each strain with the available data. Vaccination effects are also taken into account in order to study its effectiveness under scenarios with different strains.

## Model

The model used in this work is an extension of the one presented by Barreiro et al.^17^, in order to include the emergence of new virus variants at different times. The map of the country being studied is divided in a two-dimensional grid in which the dynamics occurs. We use a modified SEIRS-V model to simulate the local dynamics (see Fig. 1(A)) of infection spreading, taking place in each cell (*i, j*) of an area of a few *km*^2^, with average population density *ρ*(*i, j*). Global dynamics (Fig. 1(B)), comprising the entire country, considers geographical spread to neighboring cells and along terrestrial roads or aerial routes; governmental non-pharmaceutical interventions, and social behavior are reflected in the mobility parameters used (*v*).

**Figure 1.**
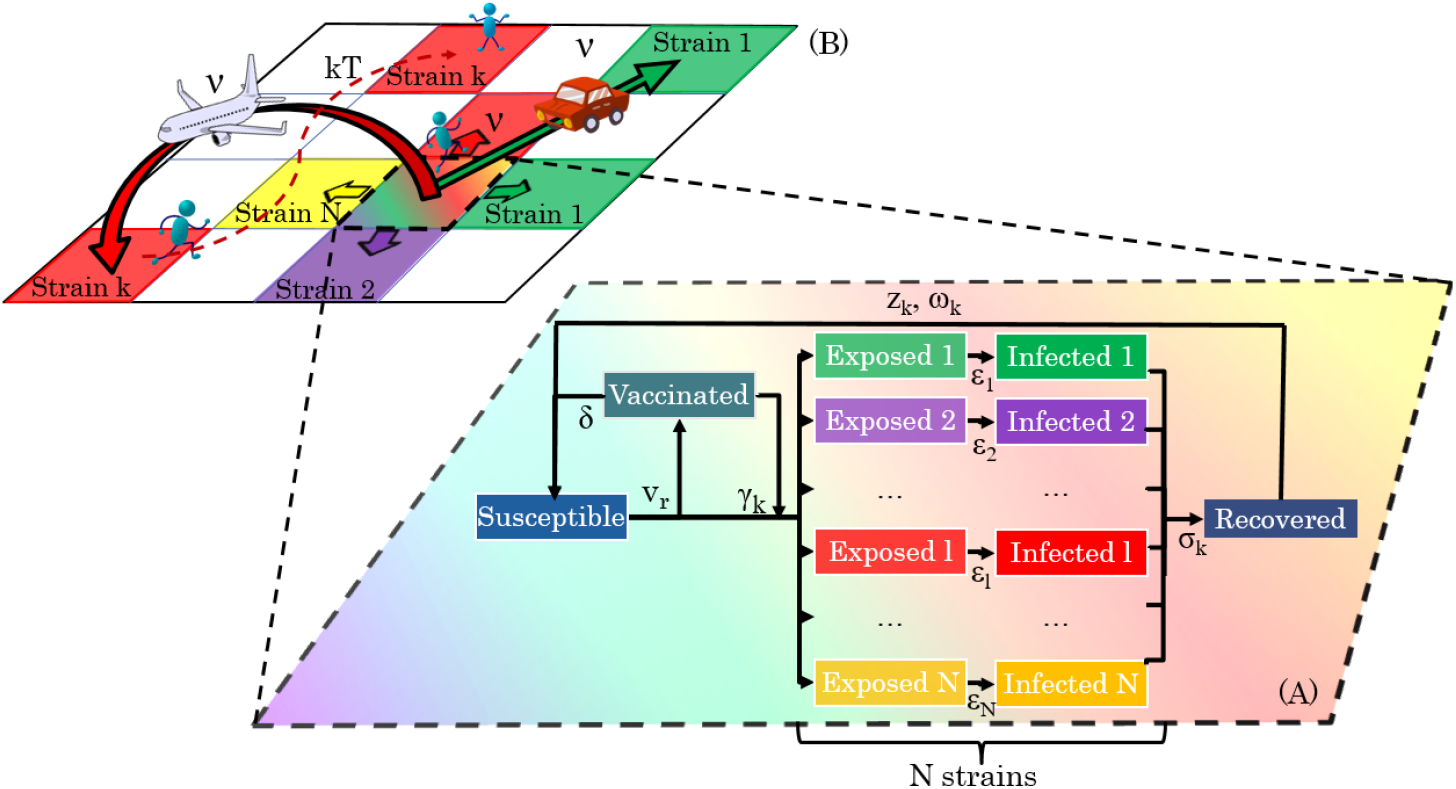
Diagram showing the geo-stochastic model scheme. (A) represents the local dynamics. Susceptible, Recovered, Vaccinated, Exposed and Infected with different strains, are taken into account. Vaccinated people can get infected with a certain variant *k* with a *γ*_*k*_ probability. The exposed, infectious and immune periods for the *k− th* strain are *ε*_*k*_, *σ*_*k*_ and *ω*_*k*_, respectively. The variables *v*_*r*_ and *δ* stand for the vaccination rate and the immunity period conferred by the vaccine. (B) The global dynamics on a geographical area, divided into a grid of cells, is followed by placing a SEIRS-V model on each one and allowing contagions between them. Three mobility processes are considered: movement to neighbor and far cells and thermal noise.

### Description of the local dynamics

The compartmental model includes: susceptible (*S*), exposed to the different variants present in the location, yet not infective (*E*_*k*_, *k* = 1,…, *N*), after *ε*_*k*_ days people become infective and can transmit the variant with which they got infected (*I*_*k*_, *k* = 1,…, *N*), they stay infective *σ*_*k*_ days and recover (*R*); acquired immunity lasts for *ω*_*k*_ days and then, a proportion *z*_*k*_ of people who survived, become susceptible again (*S*). At a specific time, vaccination designed to protect from the initial variant started, being applied at a rate *v*_*r*_; vaccinated people (*V*) stay immune for *δ* days but, with a probability *γ*_*k*_, they may be infected with variant *k* (fig 1(A)).

### Description of the global dynamic

Geographical spread (Fig. 1 (B)) is simulated by using mobility parameters: *ν*_*n*_ for movement between neighbor cells, and *ν*_*a*_ for long distance trips by airplane, car, or train. We consider that flows between large cities are greater than flows between small ones, therefore, *ν*_*a*_ is weighted by *ρ*(*i, j*)*ρ*(*m, n*) i.e. by the origin and destination population densities. All mobility parameters 0 *< ν <* 1, interpreted as the probability of traveling from one cell, in which *I*_*k*_(*i, j, t*) *≥ η*, to another cell, are modeled by a Metropolis Monte-Carlo algorithm (for details see Appendix). In the newly infected cell *S*(*m, n, t*) = 1 *−η* and *I*_*k*_(*m, n, t*) = *η*.

The epidemic reaches unexpected places because people sporadically travel to remote places, regardless of their population, these random movements may be considered as “kinetic energy” *kT* of the system. In this case we use a Monte-Carlo procedure comparing a random number with the quantity *exp*(*−kT*).

Interaction between variants can be included in our model, but until now, very few cases of co-infection with two COVID-19 variants have been reported, so we decided not to include possible interactions in this paper.

## Results

The model was applied to the case of the United Kingdom (UK). As stated above, to apply the model to a specific country data about its population density is required. The information to generate the density map was extracted from the “Global High Resolution Population Denominators Project”^18^. The main routes were obtained from Google Maps. In order to compute the geo-stochastic spreading of the virus, the map was divided into a grid of squares of 25 km^2^ (see Fig. 2).

**Figure 2.**
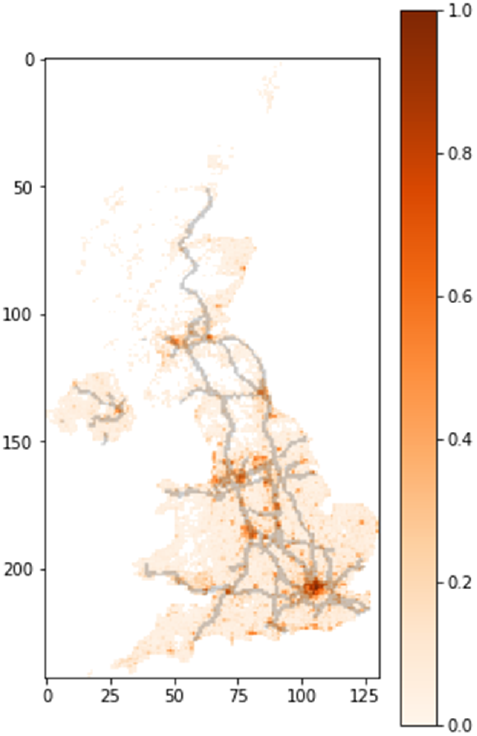
UK normalized population density and main routes map. The map is divided in a grid of squares of 25 km^2^

The dynamics of different strains of the virus is very well documented in the UK, consequently the available information suffices to fit the parameters of the model for the different variants. For simplicity, only the most abundant strains over time were considered in the adjustment. These are the EU1 strain (B. 1.177 originated in Europe), the Alpha (B.1.1.7) and the Delta (B.1.617.2). The rest of the variants, less abundant and of less interest, are grouped together under the name “other strains”. Variant information is obtained from CoVariants^19^ enabled by data from GISAID^20^. We used the same values of the time delay parameters *σ*_*k*_, *ω*_*k*_ and *z*_*k*_ for all the variants, since the model produces very good results, despite this simplification. The upper panel of Table 1 shows the values of the parameters used in order to make the calculations.

**Table 1.** Model parameters. The upper panel includes epidemiological parameters common to all strains. The lower panel shows the parameters that differ among strains.

| Parameter | Meaning | value |
| --- | --- | --- |
| $\epsilon_k$ | Period before being infectious | 1 day <sup>(a)</sup> <sup>(b)</sup> |
| $\sigma_k$ | Infectious period | 14 days <sup>(b)</sup> |
| $\omega_k$ | Immunity period | 140 days <sup>(b)</sup> |
| $z_k$ | Survival parameter | 0.99 |
| $\delta$ | Vaccination immunity period | 180/360 days |
| $\eta$ | threshold to start an infection focus | 0.00005 <sup>(c)</sup> |

<sup>(a)</sup> This value was set to 0 for the Delta variant in order to get a better fit.
<sup>(b)</sup> Values fitted and used with this model in previous works<sup>15–17</sup>.
<sup>(c)</sup> Corresponds to start an infection focus with 1 infected person in an averagely populated area.

| Parameter | other | EU1 | Alpha | Delta |
| --- | --- | --- | --- | --- |
| $\gamma_k$ | 0* | 0* | 0.01 | 0.1 / 0.3 |
| $\beta_k$ | 0.91 | 1.1 | 2.3 | 4.5 |

\* Values left as zero for simplicity, considering that when massive immunization began, Alpha was the dominant variant.

The vaccination rate *v*_*r*_ was fitted over time in order to adjust the model to the real immunization data (delayed 14 days to ensure immunity development). Vaccination and daily cases data were obtained from “Our World in Data”^2,21^ and Johns Hopkins University^1^. 66% of the population was fully vaccinated by the end of September. Taking into account the willingness to be vaccinated^2,21^ in the United Kingdom, however, it is not expected that a large population group will be vaccinated after that. For simplicity, in the model we consider that 70% of the population will be immunized.

We used the stringency index^22^ (a measure of the severity of government policies during the pandemic) as a guide to fit mobility *ν* over time. In order to reduce the amount of fitting parameters we employed the same value of *ν* for both neighbor and far mobility (the latter was weighted by the origin and destination densities). Mobility was adjusted up to August 15, 2021, leaving the same value from then on. Any change in government policies, increasing or decreasing restrictions, could produce a modification of the results. *kT* was left on a low level (0.1) since, from the beginning of the pandemic, there have been multiple measures to reduce mobility (internal restrictions and bans or quarantine for travellers entering the UK from high-risk regions)^2,22^. The parameter *β*_*k*_, which reflects how contagious a strain is, was changed to fit the actual curves for each variant. As we mentioned before, new variants could be more contagious^6,7^ and dominate over previous ones. In this sense, *β*_*k*_ determines how variants will compete and dominate over time. The lower panel on Table 1 shows the fitted *β*_*k*_ values for each strain. At the same time, some vaccines have exhibited a lower effectiveness against some variants^23^. To take this into account we used the *γ*_*k*_ parameter. Given a *k− th* strain, the higher *γ*_*k*_, the more likely that a vaccinated person will get infected by it. Some parameters such as the period of immunity conferred by the vaccine and the efficiency for some variants still need further study. We emphasize here that the immunity period that the vaccines provide, together with the inherent efficiency of the vaccines are the important factors to be considered. Therefore, we decided to propose three possible different scenarios:

- Short vaccine immunity period (*δ* = 180 days) and relatively high vaccine efficiency against delta variant (*γ*_*delta*_ = 0.1)
- Long vaccine immunity period (*δ* = 360 days) and low vaccine efficiency against delta variant (*γ*_*delta*_ = 0.3)
- Long vaccine immunity period (*δ* = 360 days) and relatively high vaccine efficiency against delta variant (*γ*_*delta*_ = 0.1)

When computing the dynamics of the model, we adjusted *β*_*k*_ and *v*_*r*_ to fit the new cases of each variant and the real vaccination rate, respectively. Fig. 3(A) shows the fitting for each variant until September 10. Adjusted values of *β*_*k*_ are shown in Table 1. The inset in the figure shows the fraction of infections with each variant. In order to fit the fast case growth of the delta variant (which cannot be explained only in terms of mobility changes) we had to use a very high *β*_*delta*_ value and a lower value of *ε*_*delta*_ (0) (see Table 1). In order to remark the evolution of the Delta variant, in this inset, we show a longer time period. From the Fig. 3(A) it is clear that at the current transmission rate this strain will dominate over the others and, unless a new, more contagious, variant appears, it will prevail in the near future. Figure 3(B) shows immunization as a function of time. In this case the vaccination rate was changed on the model to fit the actual data with a delay of 14 days. This delay was added to ensure that fully vaccinated people reach actual immunization. The dashed orange curve represent official data while the red solid line corresponds to the model immunization progress. As mentioned before, vaccination stops when 70% population is reached.

**Figure 3.**
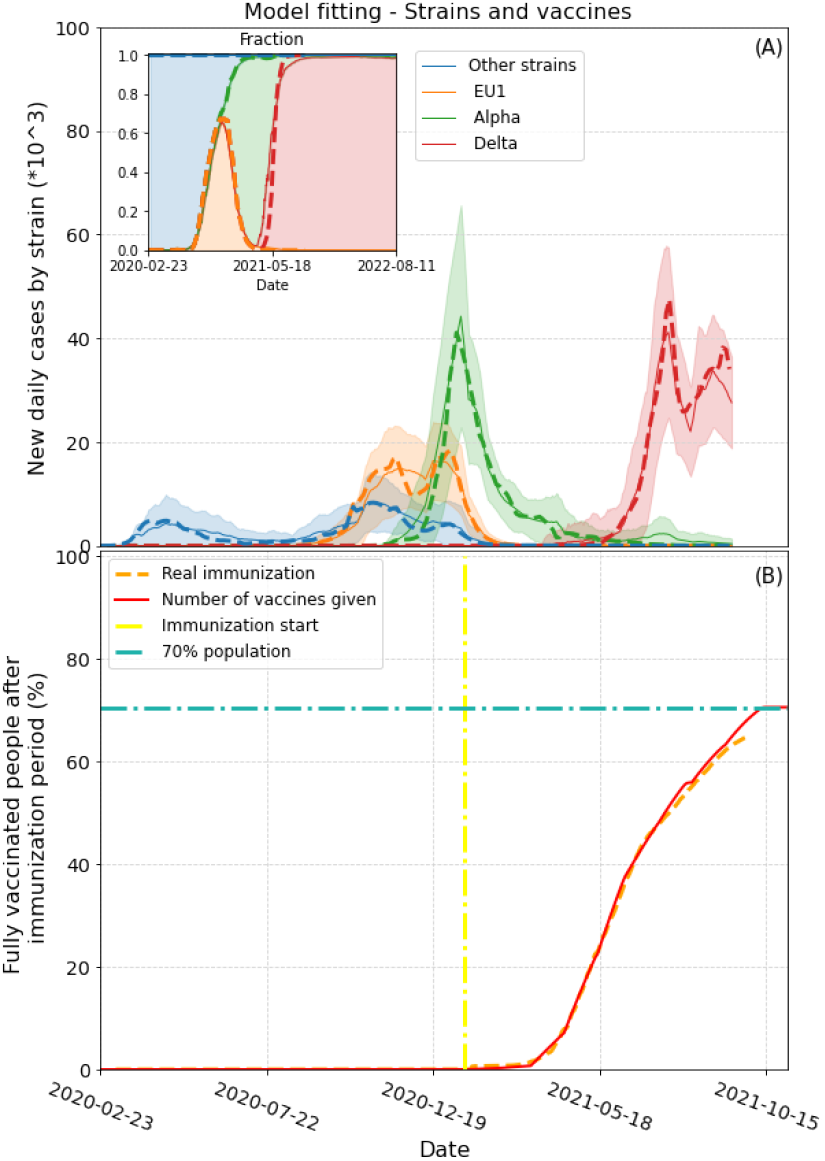
Model fitting to strain and vaccination data (A) Daily cases differentiated by strain. Dashed lines represent actual cases of each strain (scaled). Solid lines and shaded areas represent the average and standard deviations obtained from 100 runs of the model, respectively. The inset shows the fraction of cases for each variant for a longer period of time. The Delta variant is expected to prevail if no new more contagious variants appear. (B) Immunization over time. Yellow dash-dot line signals the day immunization started. Red solid line represents the percentage of fully immunized people in the model in time. The dashed orange line depicts the actual two-dose vaccination data (with 14 days delay, which is the average time needed to acquire full immunity after the second dose).

Fig. 4 shows the three scenarios mentioned above. The orange solid line represents a 7-day moving average of the new daily cases which are depicted by the blue bars. The start of the vaccination in the model is indicated with a yellow dash-dot line. The model evolution under the first scenario is represented by the green solid line. With a high efficiency vaccine, a low amount of breakthrough infections (infected while fully vaccinated) is expected in the short term, at the current mobility rate. On the other hand, a short immunity period (only 180 days) leads to a massive COVID-19 outbreak for the beginning of 2022, if no restriction measures are imposed in the future.

**Figure 4.**
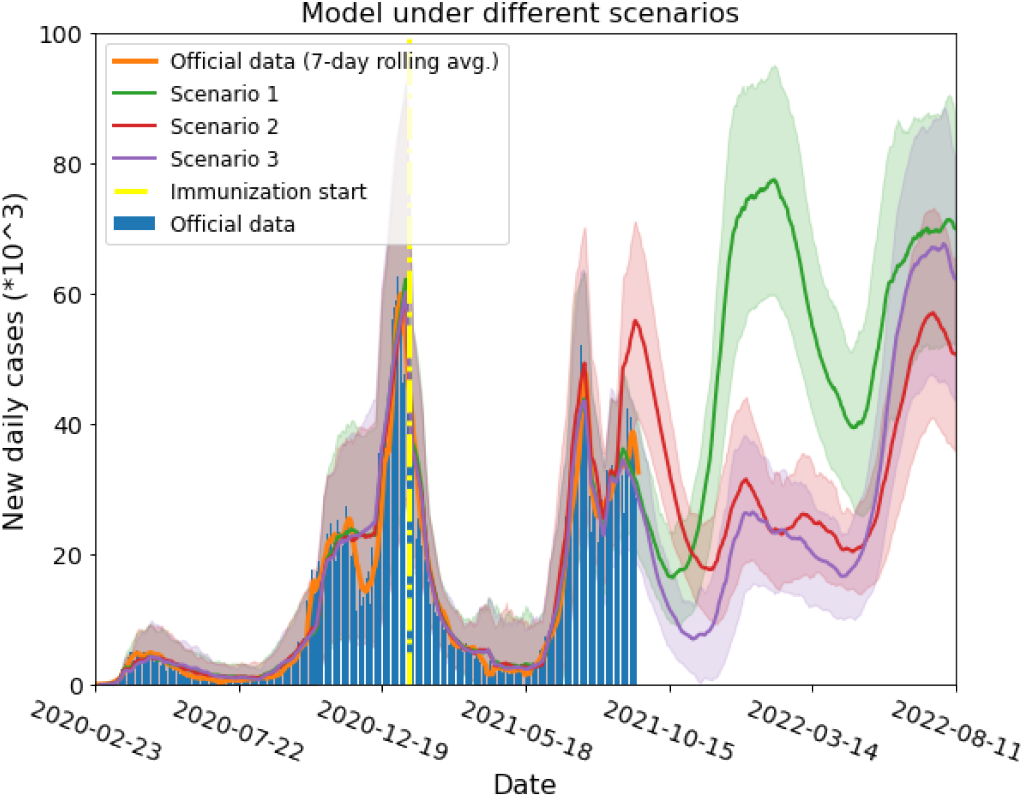
Model results for three different scenarios. Red, green and violet solid lines are the model simulation results under scenarios 1,2 and 3 (see text), respectively. Shaded areas on each color represent the standard deviation from 100 model runs for each scenario. Yellow dash-dot vertical line indicates the day when immunization started. Blue bars and solid orange lines are the actual daily cases and their 7 day rolling average, respectively. In scenario 1, a short vaccination immunity period implies a growth of the daily cases in the near future, if current mobility is sustained. Under scenario 2, people are expected to be immune for a longer time and breakthrough infections will act as antibodies boosters, prolonging the defence against the pandemic. In scenario 3 most people will be immune in the near future, lowering the number of cases, and a new wave would appear when vaccine immunity ends.

The model applied to the second scenario is shown in red. As we mentioned before, the model was conceived with the assumption that if a person presents a breakthrough infection, his/her immune system is boosted and therefore he/she will remain immune *δ* additional days after infection. This assumption is sensible, given the abundant evidence of increased antibody levels when a previously infected person is vaccinated^24–26^. This is clearly shown by the model dynamics in Fig. 4. In this scenario, while a quite large number of people have a breakthrough infection (given the a relatively low effectiveness of the vaccine against delta variant) their immune system is boosted and they stay immune for a longer period of time. Therefore, we expect a larger amount of cases in the short term, while in the long term, more people will stay immune, consistently reducing the number of cases over time. In this scenario, if the majority of the population loses their immunity, a new wave is expected in July 2022. This suggests the need of new vaccines, effective against new variants, in the years to come. Additionally, breakthrough infections could help reducing the number of cases in the long term while not increasing the amount of deaths.

The third scenario is depicted by the violet solid line in the figure. A smaller amount of cases is expected in the near future since most people are fully immunized. However, afterwards we expect a new growth in daily cases by the spring 2022. This growth is due to the loss of most people’s immunity. It is interesting to notice that, under our assumptions, the last peak in the 3rd scenario is predicted to be higher than in the 2nd one. This occurs because a lower efficacy of the vaccine implies a greater number of infections and, thus, re-immunized people. This antibody boost reduces the number of susceptible people by July 2022 lowering the height of the cases peak.

More data are needed still in order to decide which scenario is closer to the real one. However, our predictions raise questions about the proper management of the pandemic. How should vaccines be administered? Are non-pharmaceutical interventions a long-term solution? Is the pandemic here to stay? Our results suggest that new COVID-19 waves will come if highly transmissible, vaccine-resistant variants, are present. This would imply the need of new shots including strain-specific proteins. However, mass vaccination is clearly the key to reduce the appearance of new variants of concern and, consequently, the need of those specific shots. There is currently a huge debate in the world on this matter^27–29^. If vaccinated people get a mild version of the infection, COVID-19 will turn out to be a common, relatively harmless, influenza-like disease and (as shown in our model) there will be less susceptible people over time. Under this assumptions, mobility restrictions should be applied mainly to contain the spread of new variants of concern and boosters should only be given to high risk groups in the near future. Long-term booster vaccination is a different discussion to be considered, as there is no information on the duration of its immunity yet. The results of the present study seem to imply that the world’s population would probably need to be vaccinated periodically in the years to come.

## Discussion

Given the global impact of the COVID-19 pandemic, reliable models to analyze the virus propagation, including the variants of concern, are crucial to explore effective mitigation strategies. Our geo-stochastic multi-variant SEIRS-V model provides means of accurately describing the dynamics of the pandemic, as shown here. The model clearly separates the biological and social parameters. This property enables it to explain some global features observed in the daily number of worldwide cases. For instance, the surge of periodical waves is not strictly related to the appearance of new variants.These unexpected periods, have been seen in various countries with varying vaccination and social-distancing schemes. Our model can explain this global pattern, as naturally related to the biological parameters it includes, such as the immunity time of the recovered patients and the immunity time conferred by vaccines, mainly.

In the present work, we applied the model to the UK, and found that it successfully describes the dynamics of the pandemic. Notice that it was essential to fit variant-dependant epidemiological parameters like the transmission coefficients (*β*_*k*_) for the Alpha and Delta variants of the disease. We identified the epidemiological parameters which determine the dominant variant: the one possessing larger transmission coefficient (*β*) or smaller exposed period (*ε*) will eventually become dominant. Interestingly, as new variants with higher transmission coefficient appear, they quickly become the prevalent strain.

We emphasise that the model is applicable to any country with reliable data, as shown here for UK with the main variants of concern, and previously for Finland, Iceland, Mexico, Spain and Argentina^15–17^. The social parameters included in the model, as the different mobility types, allow to distinguish the effects of behavioural and cultural differences. For Spain and Argentina good fittings were obtained with the same biological parameters as in the UK (not presented here).

Furthermore, this model is useful to predict future scenarios, testing pharmaceutical and non-pharmaceutical interventions, and in particular to optimize the timing of vaccination boosters in order to minimize the appearance of new waves of the disease.

## Data Availability

Code available on request

## Appendix

### Model

#### Local dynamics

New compartments most be added to the SEIRS-V model, there are *k E* (exposed) and *k I* (infective) compartments, one compartment for each virus variant. *S* (susceptible), *R* (recovered) and *V* (vaccinated) are however, single compartments. Summation terms are used in *R* and *S* in order to include the recovered after being infected with any of the variants, that later become susceptible again. Latency periods (*ε*_*k*_), infectiousness periods (*σ*_*k*_) and immunity periods (*ω*_*k*_) are dimensionless constants expressed in units of one day.

*V* is a single compartment since available vaccines were all developed against the same variant. People become part of the vaccinated compartment only after receiving the shots recommended by pharmaceutics and having developed immunity. When vaccinated individuals get infected (with probability *γ*_*k*_), the immune system is boosted by the presence of the virus, we therefore assume they will remain immune *δ* additional days after they got infected. Products in this *V* compartment track the timing of losing immunity of this individuals.

The dynamical model equations for each cell are described by the following mathematical map,

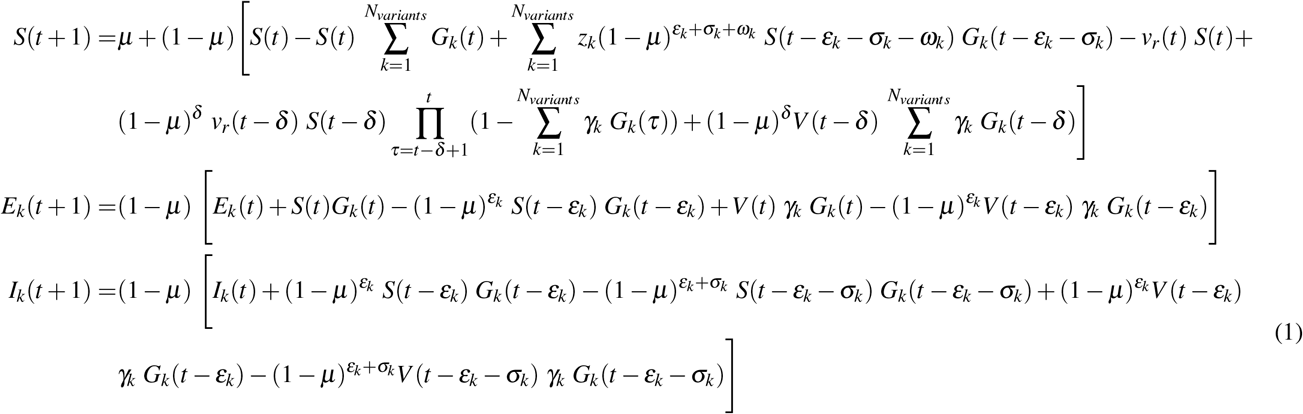

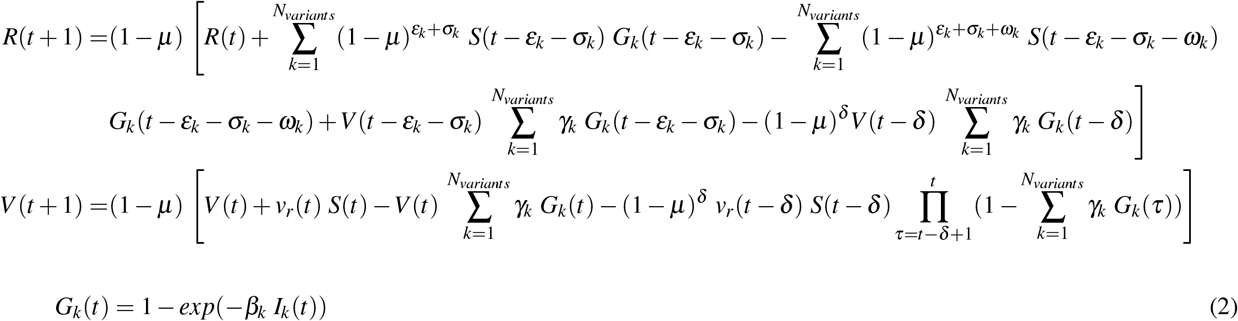

Along the simulation, population is considered to be constant,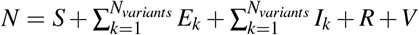; if *L* denotes life expectancy, and for the mortality we assume an exponential functional form with a constant rate *µ* = 1*/L*, birth rate should be *µN*; in each cell total population is normalized to 1. *G*_*k*_ is the Poisson based probability of becoming infected after being in contact with one or more infected people; the incidence function for each cell is *S*(*i, j, t*)*ρ*(*i, j*)*G*_*k*_(*i. j*.*t*), where *β*_*k*_ is the transmission parameter of each variant, it is a dimensionless constant which does not depend on population density or mobility of the population. Notice that in this SEIRS-V model, all time parameters remain constant, they depend only on each virus variant and immune system’s reaction, and are clearly separated from parameters affecting infection’s spread. Actual population density within each cell is considered in the incidence function as indicated before; mobility parameters are involved in geographical spread, they are modulated by social compliance of non-pharmaceutical measurements proposed by different governments.

At local level, people do not only follow a daily routine but they may also visit unpredictable places. A local mobility parameter 0 *< ν*_*L*_ *<* 1 depicts this fact, adding randomness to cell dynamics. *ν*_*L*_ is compared with a random number (*r*) from a uniform distribution, if *ν*_*L*_ *≤r* the epidemics in the cell proceeds, else no new cases are added in this cell this particular day.

#### Global dynamics

Traditional models use diffusion to model geographical spread of epidemics; we use a Metropolis Monte-Carlo algorithm to simulate mobility between cells. Although we described three different mobility parameters (*ν*_*L*_, *ν*_*n*_ and *ν*_*a*_), for simplicity we consider *ν*_*L*_ = *ν*_*n*_ = *ν*_*a*_ = *ν*, and 0 *< ν <* 1. In the case of neighbor cells, we localize cells where *I*_*k*_(*i, j, t*) *≥ η*, then if *ν≤ r*, the infections spreads to the neighboring cell (i,j+1,t) where *S*(*i, j* + 1, *t*) becomes 1 *−η* and *I*_*k*_(*i, j* + 1, *t*) = *η*; (*r* is a random number from a flat distribution, *η* is a parameter related to the infectiousness of the disease).

For long distance dispersal, the amount of travels affects the probability of spreading disease; to estimate this movement we consider that people travel more from/to large cities than from/to small ones, therefore, for long distance transmission from cell (i,j) where *I*_*k*_(*i, j, t*) *≥η* to cell (m,n), if *ρ*(*i, j*)*ρ*(*m, n*)*ν≤ r* then *S*(*m, n, t*) = 1*− η* and *I*_*k*_(*m, n, t*) = *η*.

Non-habitually trips are captured in the model by the “kinetic energy” parameter *kT*. For cells with *ρ*(*i, j*) *> T* (where T is a normalized threshold), if *exp*(*− kT*) *> r* then *S*(*i, j, t*) = 1 *−η* and *I*_*k*_(*i, j, t*) = *η* meaning that an ill person reached this cell and propagated the disease.

## Acknowledgements

RAB, MN and NLB acknowledge support from The National Autonomous University of Mexico (UNAM) and Alianza UCMX of the University of California (UC), through the project included in the Special Call for Binational Collaborative Projects addressing COVID-19. RAB was financially supported by Conacyt through project 283279 and PAPIIT, UNAM project IG200121. MN was partially supported by CONICET. NLB and PGB acknowledge support from project 03 NAC 009/21 from CITEDEF. We acknowledge the researchers and laboratories responsible for obtaining the specimens, generating the genetic sequence and metadata and sharing via the GISAID Initiative.

## Author contributions statement

N.L.B., T.G. and R.A.B. conceived the idea. N.L.B. proposed the model, and wrote the program in python. N.L.B. created the maps, did the fitting and conducted the simulations. All authors discussed the results, wrote and reviewed the manuscript.

